# SARS-Coronavirus-2 nucleocapsid protein measured in blood using a Simoa ultra-sensitive immunoassay differentiates COVID-19 infection with high clinical sensitivity

**DOI:** 10.1101/2020.08.14.20175356

**Authors:** Dandan Shan, Joseph M Johnson, Syrena C. Fernandes, Muriel Mendes, Hannah Suib, Marcella Holdridge, Elaine M Burke, Katie Beauregard, Ying Zhang, Megan Cleary, Samantha Xu, Xiao Yao, Purvish Patel, Tatiana Plavina, David Wilson, Lei Chang, Kim M Kaiser, Jacob Natterman, Susanne V Schmidt, Eicke Latz, Kevin Hrusovsky, Dawn Mattoon, Andrew J. Ball

## Abstract

The COVID-19 pandemic continues to have an unprecedented impact on societies and economies worldwide. Despite rapid advances in diagnostic test development and scale-up, there remains an ongoing need for SARS-CoV-2 tests which are highly sensitive, specific, minimally invasive, cost-effective and scalable for broad testing and surveillance. Here we report development of a highly sensitive single molecule array (Simoa) immunoassay on the automated HD-X platform for the detection of SARS-CoV-2 Nucleocapsid protein (N-protein) in venous and capillary blood (fingerstick). In pre-pandemic and clinical sample sets, the assay has 100% specificity and 97.4% sensitivity for serum / plasma samples. The limit of detection (LoD) estimated by titration of inactivated SARS-CoV-2 virus is 0.2 pg/ml, corresponding to 0.05 Median Tissue Culture Infectious Dose (TCID50) per ml, > 2000 times more sensitive than current EUA approved antigen tests. No cross-reactivity to other common respiratory viruses, including hCoV229E, hCoVOC43, hCoVNL63, Influenza A or Influenza B, was observed. We detected elevated N-protein concentrations in symptomatic, asymptomatic, and pre-symptomatic PCR+ individuals using capillary blood from a finger-stick collection device. The Simoa SARS-CoV-2 N-protein assay has the potential to detect COVID-19 infection via antigen in blood with performance characteristics similar to or better than molecular tests, while also enabling at home and point of care sample collection.

**One Sentence Summary:** SARS-CoV-2 nucleocapsid protein (N-protein) measured in serum, plasma, and dried blood spots (DBS) via ultrasensitive immunoassay can be used to differentiate PCR+ from PCR- patients, even if asymptomatic.

## Introduction

In November 2019, the SARS-CoV-2 (severe acute respiratory syndrome coronavirus-2) emerged in Wuhan, China and since has caused a worldwide pandemic ^1^. To date, the USFDA has granted Emergency Use Authorization (EUA) to three types of SARS-CoV-2 assays: molecular testing or PCR, antibody testing or serology, and antigen testing^2^. Molecular testing for viral RNA is the primary diagnostic modality for active infection, while serology measures anti-SARS-CoV2 antibodies post-infection ^3,4^. Although RT-PCR-based molecular testing for viral RNA in respiratory specimens is the primary diagnostic tool for active infection, concerns have been raised about the risk of false negative results associated with the use of nasal and nasopharyngeal swabs ^5^. This is especially true in the days before symptom onset; Kucirka et al. have found the probability of a false negative result in an infected person to decrease from 100% on day 1 post-infection to 67% on day 4. On day 5, the median time for symptoms to appear, molecular tests still had a 38% probability of producing a false negative result and declined no further than 20% in the days that followed, when the infection should be most detectable^6^. Furthermore, the complexity, cost, supply chain challenges, and relatively lower throughput of RT-PCR results are disadvantages toward fulfilling large-scale testing required to enable societies to re-open^7^.

Antigen detection by immunoassay has the potential advantages of a simpler workflow, faster turn-around time, lower cost, and with a supply chain diversified from PCR. However, currently available antigen tests are generally less sensitive than PCR, for example one lateral flow assay has been reported to have percent positive agreement (PPA) with qRT-PCR of only 24 to 30%^8,9^. Two EUA cleared antigen tests have claimed sample types of nasopharyngeal or nasal swabs with 96.7% and 84% PPA with PCR and should greatly enhance diagnostic capacity, but they are still subject to the same sampling challenges associated with nasal or nasopharyngeal swabs and less analytical sensitivity relative to PCR ^10,11^.

SARS-CoV-2 infections can present unusual peripheral symptoms, such as stroke, heart attack, kidney damage, neurological symptoms, and COVID-toe. These clinical manifestations suggest that this respiratory virus can migrate from the lungs into the bloodstream. Mehra et al. first described evidence of SARS-CoV-2 peripheral involvement during post-mortem histological examination of effected tissues, including electron microscopy images of viral inclusion structures in endothelial cells ^12^. It was hypothesized that SARS-CoV-2 infection may facilitate the induction of endothelitis in multiple organs as a direct consequence of viral involvement. Wölfel et al. reported that SARS-Cov-2 virus was not detectable in blood using molecular diagnostic techniques^13^, but additional later studies have found evidence that plasma viremia may play a significant role in disease course and that viral loads in plasma may predict risk of death^14^.

Recently, Ogata et al. measured SARS-CoV-2 antigens (S1 antigen, spike antigen, N-protein) in venous blood for the first time^15^. They hypothesized that detection of viral antigen could be used to stratify patients between mild and severe cases, but that asymptomatic or mild cases would not have measurable levels. If true, this would be a distinct difference between SARS-CoV-2 and SARS-CoV, as patients of the latter had measurable levels of N-protein in blood up to 3 weeks after symptom onset, and measurement of N-protein had 94% PPA up to 5 days compared to PCR ^16^.

We theorized that by leveraging the exceptional sensitivity of Single Molecule Array (Simoa) immunoassay technology, we could detect and quantitate SARS-CoV-2 antigen directly in serum and plasma from venous collection and capillary blood acquired by commercially available finger-stick collection devices. Here we report the development of a blood-based assay for SARS-CoV-2 N-protein that potentially shows detection of clinically significant viral loads in active and pre-symptomatic COVID-19 infections, avoiding the use of swabs and the need to sample nasopharyngeal or nasal fluids.

## Materials and Methods

### Samples

Healthy pre-pandemic serum and plasma samples (collected before December 2019) were obtained from BiolVT (Westbury, NY). Commercially sourced serum and plasma samples from COVID-19 positive donors, as demonstrated by positive RT-PCR test, were obtained from BiolVT and from Boca Biolistics (Pompano Beach, FL; hereafter ‘BocaBio’). Samples were collected between April 06 and June 17, 2020. RT-PCR was performed between March 06 and June 12, 2020. Plasma samples from hospitalized COVID-19 patients, as demonstrated by positive RT-PCR test, were provided by Drs. Jacob Nattermann, University of Bonn, Germany. Samples were collected between March 30 and April 22, 2020. RT-PCR was performed between March 30 and April 15, 2020. The study was approved by the Institutional Review board of the University Hospital Bonn (134/20). Patients were included after providing written informed consent. In COVID-19 patients who were not able to consent at the time of study enrollment, consent was obtained after recovery. Dried blood microsamples were collected using Mitra® Devices (Neoteryx, Torrance, CA) from staff and residents of CT Baptist Care Homes Inc. (CTCH cohort). COVID-19 status of each donor was determined by RT-PCR test and DBS samples were collected at two time points, one week apart, for measurement of N-protein and IgG levels by Simoa. Gamma-inactivated SARS-CoV-2 virus was obtained from BEI (beiresources.org), heat-inactivated SARS-CoV-2 and microbial specimens for cross-reactivity testing were obtained from ZeptoMetrix. (zeptometrix.com).

### Assay Development

In Research Use Only (RUO) products Single Molecule Array (Simoa) technology offers sensitivity on average 1000-fold greater than traditional immunoassays ^17,18^. In brief, the technology involves performing a paramagnetic microbead–based sandwich ELISA, followed by isolation of individual capture beads in arrays of femtoliter-sized reaction wells. Singulation of capture beads within microwells permits buildup of fluorescent product from an enzyme label, so that signal from a single immunocomplex can be detected with a CCD camera in 30 seconds. At very low analyte concentrations, Poisson statistics dictate that bead-containing microwells in the array will contain either a single labeled analyte molecule or no analyte molecules, resulting in a digital signal of either "active" or "inactive" wells. Data collection involves counting active wells corresponding to single enzyme labels. At higher analyte concentrations, digital measurements transition to analog measurements of total fluorescence intensity. Simoa data are reported as Average Enzymes per Bead (AEB). It is widely used in the field of neurodegenerative disease and recently, for the measurement of SARS-CoV-2-associated biomarkers ^19,20^. It has also been demonstrated to rival the sensitivity of PCR for monitoring HIV infection through measurement of the p24 capsid protein in blood ^21,22^.

#### SARS-CoV-2 N-protein Assay

Antibodies and antigens were obtained from commercial sources. Eight different antibodies and five antigens were screened, resulting in more than 60 different test configurations. The antibody and antigen combination that produced the best signal / background ratio for both calibrator and positive samples was selected. Diluent formulations, detector antibody and Streptavidin–β-Galactosidase concentrations were then optimized, as well as assay protocols (2-step vs 3-step; incubation times). A phosphate-based sample diluent was selected with EDTA to inhibit proteases, heterophilic blocker and a detergent to help de-envelope and inactivate virus particles.

#### SARS-CoV-2 IgG Assay

An assay was developed to monitor the serological response of IgG to the full-spike of SARS-CoV-2. This assay has been submitted for EUA clearance (USFDA application EUA20164); verification and validation details are planned to be released in product-specific validation reports and instructions for use upon product launch.

## Assay Verification

<u>N-protein Assay.</u> The assay was verified by testing 6 runs over 3 days over 2 lots, for a combined total of 12 runs. Verified characteristics include precision, ad-mixture linearity, spike recovery, limit of blank (LoB), limit of detection (LoD), and limit of quantification (LoQ) for serum, K2EDTA plasma, and dried blood spots (Supplementary Figure S1 and Table S1). Precision was determined using 2 diluent-based controls and 3 matrix based spiked samples. Limit of detection (LoD) was determined with gamma-inactivated SARS-CoV-2 virus diluted 6e6 fold into a serum from a negative donor, using stock with a TCID_50_ of 2.8e5 per mL (final TCID_50_ = 0.05 per mL), and testing 36 replicates over 3 days and 2 different assay lots. Admixture linearity was demonstrated using negative matrix spiked with heat-inactivated virus, and then mixed in varying ratios with a separate non-spiked matrix to create ten levels. We established a preliminary clinical cutoff by measuring N-protein levels in four SARS-CoV-2 negative cohorts: 1) pre-pandemic serum / plasma (N=100); 2) a panel of plasma samples sero-negative for SARS-CoV-2 negative and sero-positive for common respiratory infections (N=36, Supplementary Table S2); 3) a panel of serum samples sero-negative for SARS-CoV-2 and sero-positive for other common coronaviruses (N=31, Supplementary Table S3); 4) PCR- DBS samples from CTCH (N=9). Data is shown in Supplementary Figure S2.

### Sample Types

Serum, K_2_EDTA plasma, and dried blood spots were used in the analyses. Serum and plasma were collected by normal processing methods, and stored frozen at -80C before analysis. Serum and plasma samples were diluted 4-fold into assay diluent on the HD-X instrument before measurement. Dried blood spots (DBS) were collected using Mitra collection kits from Neoteryx according to standard protocols (https://www.neoteryx.com/home-blood-blood-collection-kits-dried-capillary-blood). Tips absorb 20 μl of whole blood and are then allowed to dry for at least 16 hours in a pouch with dessicant. Tips are extracted into 250 μl of assay diluent with shaking at 400 rpm overnight at 2 - 8°C, resulting in a 12.5-fold sample dilution. All sample results have been corrected for dilution factors, to represent the concentration within the sample matrix.

### Sample Matrix Correlation

To correlate serum and plasma matrices, matched samples from PCR+ donors were measured with the N-Protein assay. N-protein levels correlated between matrices with a slope of 1.12 and an R^2^ of 0.995 (Supplementary Figure S3). To verify the recovery of N-protein from the Mitra tips, whole blood was collected into K_2_EDTA tubes, spiked with recombinant N-protein and then processed into either plasma or DBS. N-protein levels were measured in both sample types. N-protein levels correlate between matrices with R^2^ = 0.993 and a slope of 1.97. The concentration in DBS was approximately ½ of that in plasma, as expected due to the excluded volume of hematocrit which is separated from plasma (Supplementary Figure S4).

### DTT treatment of plasma samples

To determine whether seroconversion and antigen-masking by immunoglobulins plays a role in the decrease of N-protein signal, samples were treated with 10 mM DTT at 37°C for 15 minutes. To demonstrate the effectiveness of this treatment the following experiment was conducted: 1) negative serum was spiked with N-protein and measured on the N-protein assay; 2) a 500x concentration of anti-N-protein antibody was added and the sample was measured, resulting in a 60% decrease in antigen; 3) the sample spiked with both antigen and antibody was treated with DTT according to the protocol above and measured, resulting in a 75% rescue of antigen signal (Supplementary Figure S5).

### Cross reactivity studies

Cultured and inactivated pathogens were spiked into negative serum samples to attain 10^5^ TCID50 per ml, using a minimum of 4x dilution of viral stock into serum. Some virus cultures had insufficiently high stock titer to achieve 10^5^ TCID50 per ml, and these viruses were tested at the highest titer possible after a 4x dilution into serum. No cross-reactivity was observed, as detailed in Supplementary Table S4.

## Results and Discussion

To determine the clinical utility of the N-protein assay for serum and plasma, we measured PCR+ samples from BocaBio and the University of Bonn, and pre-pandemic samples from BioIVT. Figure 1 panel A represents only "first-draw" samples, in which every data point represents a unique donor. This use-case is appropriate for a test that is intended to screen novel patients as positive or negative^23^. Our preliminary cutoff of 0.9 pg/mL (dashed line) confers a clinical sensitivity of 97.6% (37/38 positives >0.9 pg/ml) and clinical specificity of 100% (100/100 negatives < 0.9 pg/ml).

**Figure 1.**
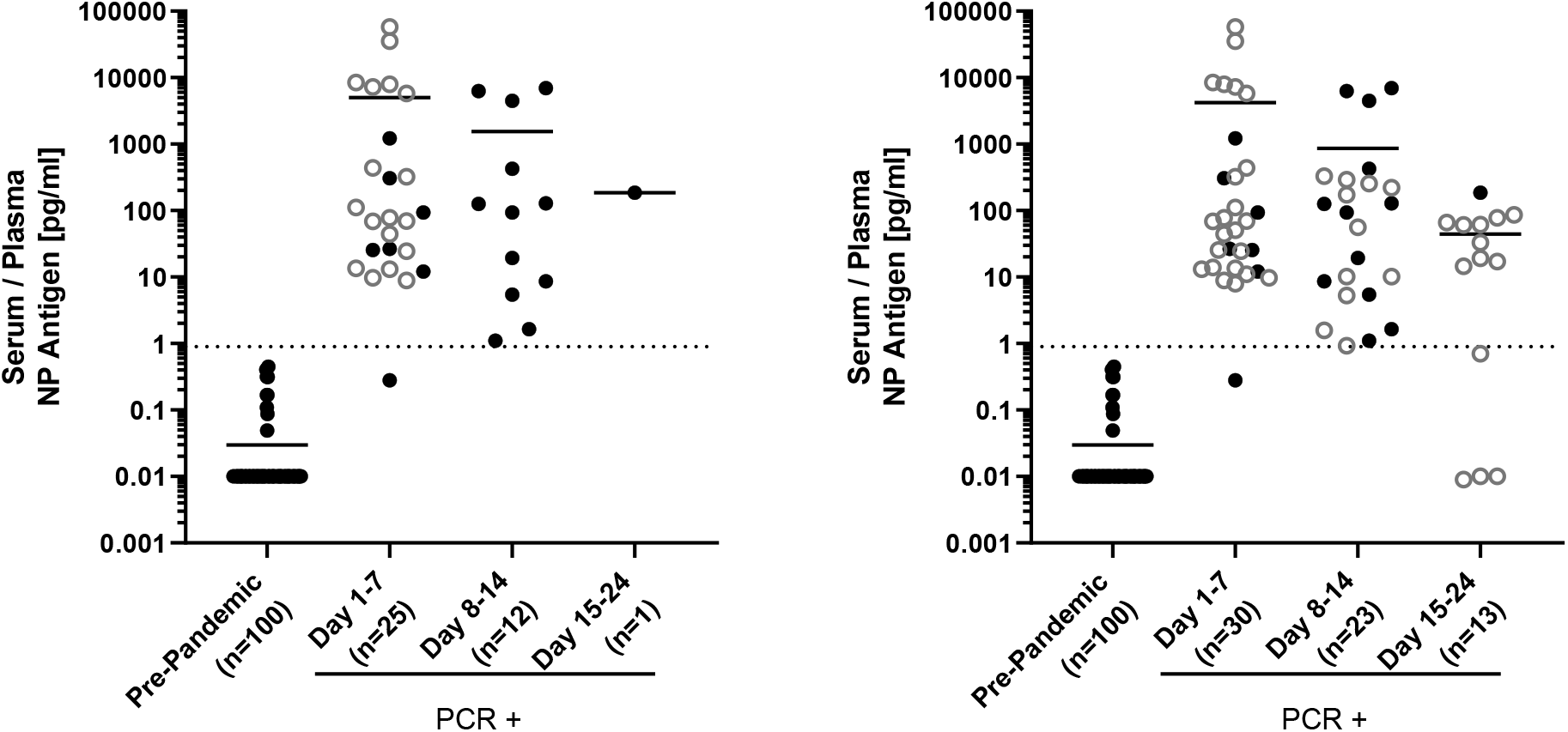
Simoa SARS-CoV-2 N Protein measurements differentiate pre-pandemic from PCR+ donors in serum and plasma. **Panel A**. Pre-pandemic sera from BioIVT (closed symbols), PCR+ sera from BocaBio (closed symbols) and the plasma samples from U. Bonn (open symbols). PCR+ samples are binned chronologically according to day-from-symptom or, if asymptomatic, day- from-PCR (BocaBio) and day-from-hospitalization / PCR (U. Bonn). Each point represents a unique donor. **Panel B**. Measurements from all samples, including multiple longitudinal draws from the U. Bonn patients.

We binned the samples by day, and in Figure 1 panel B we include multiple timepoints from longitudinal donors (Univ. Bonn) to develop an initial temporal profile of the viral antigen in blood. Using an immunoassay for SARS-CoV N-protein, Che et al. observed clinical sensitivity of 94%, 78% and 27% for blood samples within days 1-5, 6-10 and 11-20 of symptom onset ^16^. Our data shows similar performance,albeit with enhanced sensitivity, notably after the 1^st^ week of infection. This suggests the possibility that an ultrasensitive antigen assay could expand the diagnostic window beyond that addressable by the current EUA approved antigen assays that claim clinical sensitivity only within the first 5 to 7 days after onset of symptoms ^10,11^. To determine this, future studies will need to test a sample cohort with well-defined clinical characteristics, in which the onset of infection and symptom are accurately known.

We also measured anti-SAR-CoV-2 specific IgG in the longitudinal samples from the U. Bonn cohort (Figure 2). N-protein concentration in plasma was observed to decrease over time with a concurrent increase in anti-SARS-CoV2 IgG levels. By normalizing patient responses and using a four-parameter logistic regression to the average response, we find N-protein clearance to occur at 15.6 days and IgG plateau at 7.7 days after hospitalization. Several of these patients had already undergone seroconversion prior to first collection; given that seroconversion for SARS-CoV-2 can occur between day 7 to 13 post-symptom^15,24^, we estimate that N-protein clearance occurs between days 22 to 28 and IgG plateau between days 14-20 post-symptom, similar to timelines observed for SARS ^16^. Ogata et al. also observed similar timelines for SARS-CoV-2, although in their study N-protein was generally not detectable once IgG levels had stabilized^15^, whereas we observed a window of approximately 7 days between IgG plateau and N-protein clearance during which both biomarkers are quantifiable. These data suggest the value of conducting additional studies to further characterize the relationship between IgG and N-protein levels in a larger sample set.

**Figure 2.**
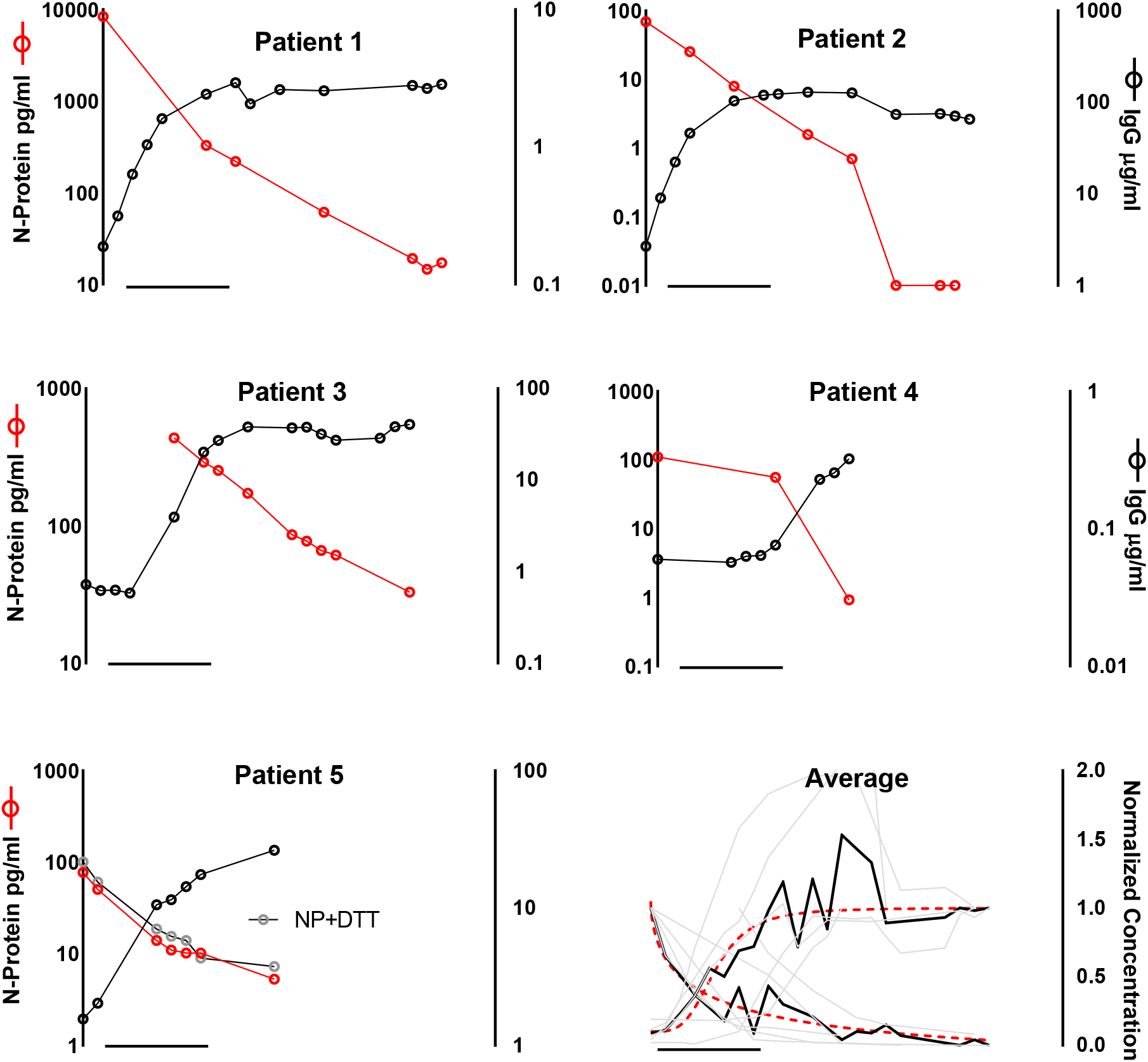
Plasma levels of anti-SARS-CoV-2 IgG increases concurrently with decreasing N-protein levels. Longitudinal samples from five patients in the U. Bonn cohort are shown. Samples from patient 5 were tested with and without DTT treatment (bottom left panel). Four-parameter logistic regression to the average, normalized concentration of N-protein and IgG (bottom right panel).

To determine whether seroconversion and antigen-masking by immunoglobulins plays a role in the decrease of N-protein signal, we treated longitudinal samples from patient 5 with DTT to unmask potential antigen-antibody complexes. We observed a modest increase in N-protein levels of 27% after treatment on average (Figure 2 Patient 5). Considering the overall decrease of >1400% over the entire time-course, we hypothesize that antigen-masking plays a negligible role, and that instead N-protein levels decrease due to clearance from the blood.

To allow at-home or point-of-care collection of blood samples, we tested dried blood spots (DBS) collected with Mitra^®^ tips (Neoteryx.com). These devices absorb 20 μl of capillary blood from a finger-stick, and users may subsequently store and ship them without cold-chain requirements. We measured N-protein levels in DBS patient samples collected in the presence of active COVID-19 infections using the Mitra devices (CTCH cohort). This long-term care facility has established a practice of testing residents and staff for COVID-19 weekly using an authorized molecular test. This enabled a comparison of the performance of the Simoa SARS-CoV-2 N-Protein Assay against the gold-standard of PCR in the context of active and on-going COVID-19 infections; relative days of collection are shown in Table 1.

**Table 1.**
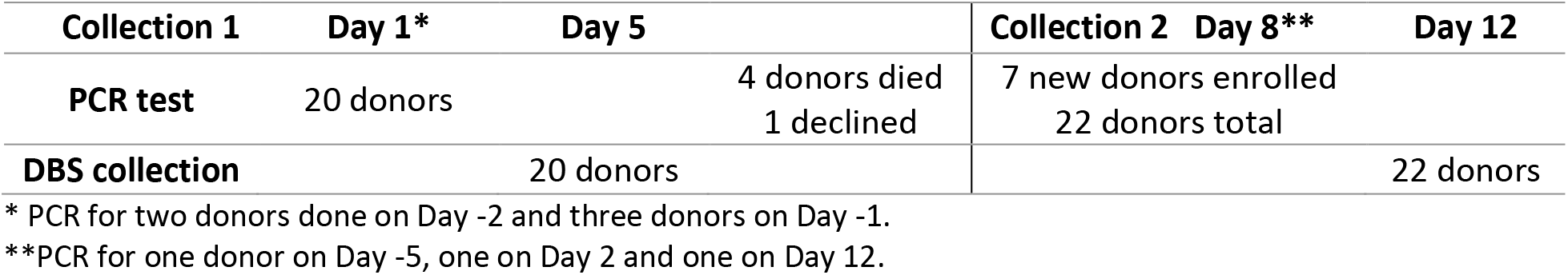
Sampling and testing timeline in CTCH study

In Figure 3 panel A we show N-protein levels for both collections from CTCH, with connecting lines denoting changes in individual donor levels from week 1 to week 2. This data demonstrates 100% sensitivity and specificity of the Simoa N-protein assay compared to PCR, and notably the Simoa N-protein assay identified COVID-19 positive status for four donors that exhibited no symptoms over the course of infection (asymptomatic) and five donors that developed symptoms after sample collection (pre-symptomatic).

The time course of donor 12 in particular illustrated the ability of N-Protein in capillary blood to diagnose COVID-19 before symptom onset: enrolled as a negative control and tested PCR- on day 1; DBS sampled on day 5 showed elevated levels of N-Protein (first collection) before symptom onset; confirmed positive with PCR testing on day 7; symptoms developed on day 8; by day 15 recovered. Donor 12 may represent a false-negative PCR result that was detected using the N-protein assay, though PCR test and DBS collection were five days apart; future studies will aim to address this question through direct comparison of clinical sensitivity of PCR and the Simoa SARS-CoV-2 N-Protein assay on samples collected concurrently. Regardless, donor 12 exemplifies the ability of the Simoa SARS-CoV-2 N-Protein assay to detect pre-symptomatic COVID-19 infection.

False negative PCR results represent a significant challenge in the COVID-19 pandemic^5^. Kucirka et al. report the highest probability of PCR false-negative results before symptom onset, with the false-negative rate decreasing from 100% to 67% in the first four days post-infection. On day 5, the median time for symptom-onset, the probability of a false negative result in a PCR-based molecular test was still 38% ^6 26^. Compounding the problem of poor clinical discrimination in pre-symptomatic patients, He et al. observed the highest viral load in throat swabs at time of symptom onset, and inferred that infectiousness will peak at or before symptom onset^25^. In this context, the ability of the Simoa SARS-CoV-2 N-Protein assay to detect pre-symptomatic individuals could be particularly important.

**Figure 3.**
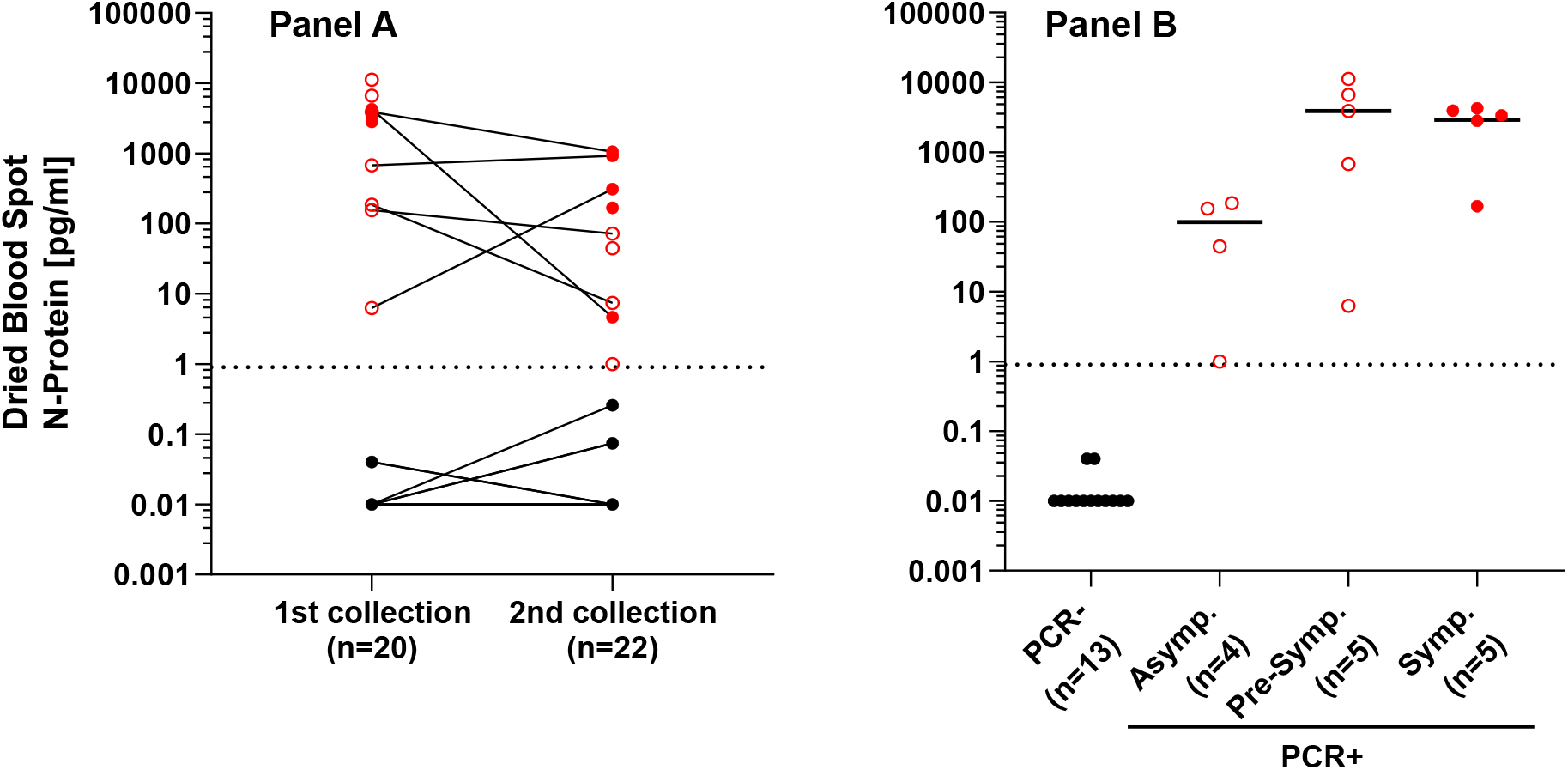
SARS-CoV-2 N-protein levels measured in capillary blood (dried blood spots (DBS)) from CTCH residents and staff confirm PCR results. **Panel A**: PCR- samples are denoted in black (•), PCR+ in red 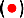, and PCR+ asymptomatic or pre-symptomatic in open red 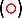 with lines connecting samples from the same donor over two collections. **Panel B**: Donors grouped into PCR-, asymptomatic PCR+, pre-symptomatic PCR+ and symptomatic PCR+, as noted at time of confirmatory PCR. Only the first collection point is represented for each donor.

In the CTCH cohort, 8 of the 14 PCR+ donors presented without symptoms even with elevated levels of N-protein and in Figure 3 panel B, we separated donors into four groups: PCR-; asymptomatic PCR+ that did not show symptoms at any point during infection; pre-symptomatic PCR+ that did not show symptoms at the first collection but developed symptoms by the second collection; and symptomatic PCR+ that presented with symptoms at the first collection. Fully asymptomatic donors have a lower median level of N-protein; however we do not see different levels of N-protein between pre-symptomatic or symptomatic donors. Ogata et al. suggested that viral antigen would only present in blood in severe or late-stage cases, however our data suggest that some mechanism exists for viral antigen to transfer to blood even in early and asymptomatic cases ^15^. Che et al. reported similar trends for SARS-CoV patients, who had a higher positive detection rate of N-protein in serum samples within the first 10 days of infection than that detected by RT-PCR in respiratory samples, an observation hypothesized to be associated in part with respiratory specimen collection variables leading to false negatives ^16^.

In Figure 4 panel A we have ranked CTCH PCR+ donors by N-protein level, and color-coded results according to disease outcome: deceased; not recovered at collection 2; or recovered at collection 2. In this limited sample set, we observe a trend of worse clinical outcome associated with higher N-protein level. Ogata et al. have observed a similar trend for viral antigen in blood^15^, and Fajnzylber et al. reported that viral-RNA load is associated with increased disease severity and mortality^14^.

**Figure 4.**
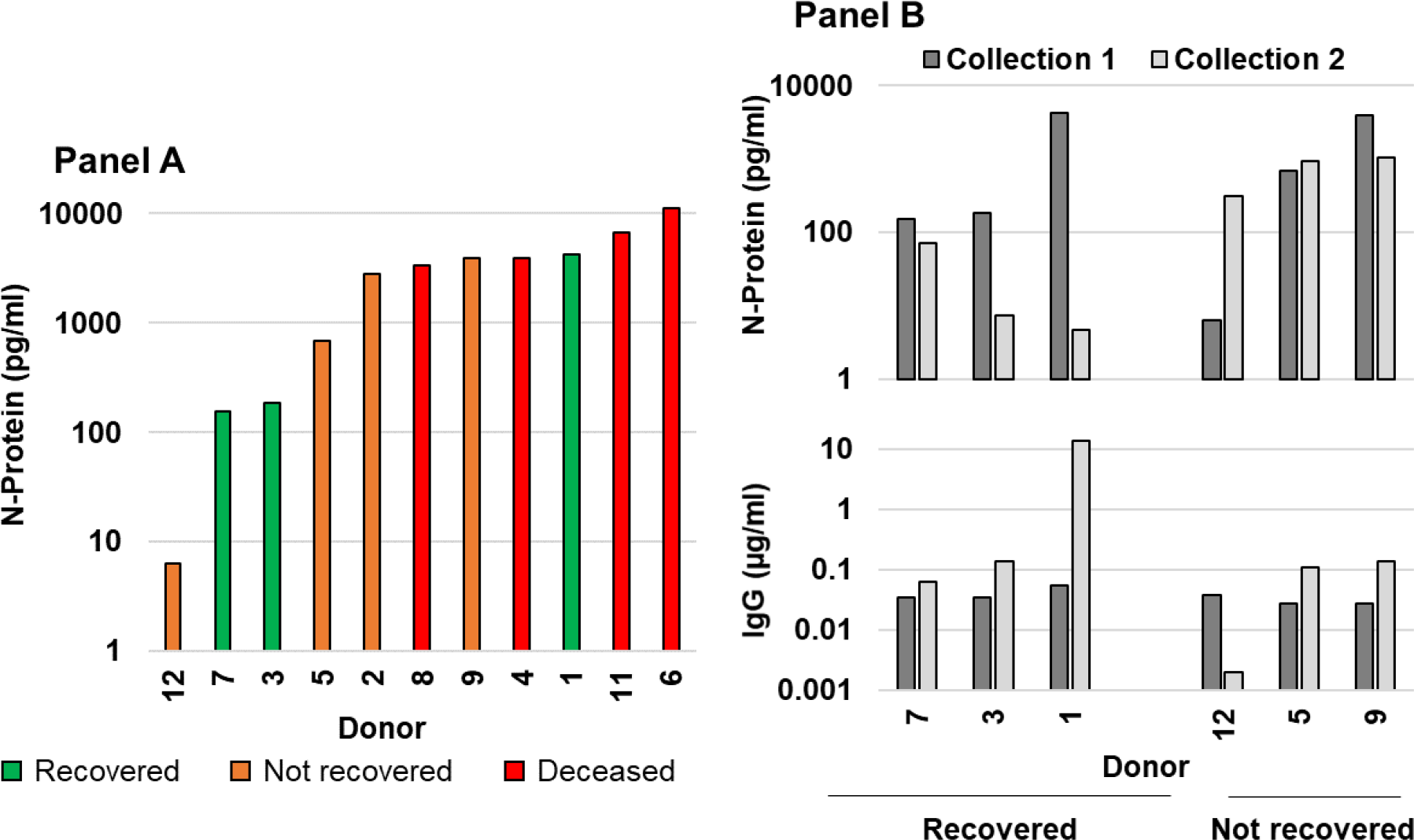
Comparison of N-protein levels from DBS with clinical severity indicators in CTCH cohort. (A) SARS-CoV-2 N protein concentrations at the initial sample collections. (B) N protein clearance after one week, and comparison to IgG levels.

In Figure 4 panel B, we have grouped donors into recovered (n=4) and not recovered (n=2) and display N-protein and IgG levels for both collections. Average N-protein level decreases 10-fold (1143 to 98 pg/ml) for recovered donors across both collection dates, contrasted with a higher starting average and more moderate decrease of 2-fold (2287 to 988 pg/ml) for not recovered donors. We measured low IgG levels for all donors at collection 1, suggesting that seroconversion had not yet occurred. At collection 2 we detected a slight IgG increase for some donors, but a large increase only for donor 1. This donor also had a concomitant, large decrease in N-protein, and was the only donor with high N-protein levels to recover by the 2^nd^ collection. Serological assessment may complement the N-protein assay and help stratify outcomes of severe cases. All data for the CTCH cohort is shown in Supplementary Table S5.

## Conclusion

In summary, we have developed a blood-based assay for SARS-CoV-2 N-Protein and our studies demonstrate detection of clinically significant viral loads in active, pre-symptomatic and asymptomatic COVID-19 infections, using sample collection methods that avoid swabs and the need to sample nasopharyngeal or nasal fluids. Based on testing performed to date, we estimate a clinical sensitivity of 97.4% in serum / plasma using two PCR+ cohorts and clinical specificity of 100% using a cohort of 100 pre-pandemic samples. We see no cross-reactivity to other common respiratory viruses, including hCoV229E, hCoVOC43, hCoVNL63, Influenza A or Influenza B. Using titers of gamma-inactivated virus we estimate the limit of detection (LoD) of our assay to be 0.05 TCID_50_, > 2000 times more sensitive than current antigen tests with EUA approval for use in nasal swabs^10,11^.

We have demonstrated detection in capillary blood using the Neoteryx Mitra^®^ dried blood spot (DBS) collection device, which enables at-home and point-of-care sample collection. Using DBS samples, we successfully monitored disease status of staff and residents in the presence of active COVID-19 infections with clinical sensitivity comparable to molecular testing in our preliminary experiments. Higher concentrations of N-protein associated with increased disease severity and mortality, and vice-versa clearance of the antigen associated with greater recoverability.

We plan further studies to validate the ability of the SARS-CoV-2 N-Protein assay to diagnose COVID-19 and determine if it has comparable or better sensitivity than molecular testing, including studies with larger, prospective cohorts with better characterized clinical symptoms and timelines. In particular, we will attempt to conduct trials with well-defined onset of infection to determine the window of effectiveness of the SARS-CoV-2 N-Protein assay, which may be able to diagnose both earlier than molecular testing (pre-symptomatic infection) and later than current EUA cleared antigen tests (beyond one week post-symptom).

The SARS-CoV-2 antigen assay has the potential to be available for widespread deployment through minimally invasive remote and home sample collection and utilization of the fully automated HD-X immunoassay platform. It is expected that this SARS-CoV-2 product candidate antigen assay may provide a new, orthogonal method for early detection of SARS-CoV-2 infection that can significantly and rapidly augment the accuracy and availability of the SARS-CoV-2 testing arsenal.

Safe Harbor Statement: CAUTION: Investigational device. Limited by federal law to investigational use. Not available for sale.

## Data Availability

All data will be made available upon written request submitted to the corresponding author.

## Acknowledgements

Special thanks to Patricia Morse and the residents and staff at CT Baptist Care Homes Inc as well as the University of Bonn, for collecting and providing specimens used in this study. This work was partly funded through a Rapid Acceleration of Diagnostics (RADx) grant from the National Institutes of Health, awarded through the University of Massachusetts Medical School.

---

**Figure S1.**
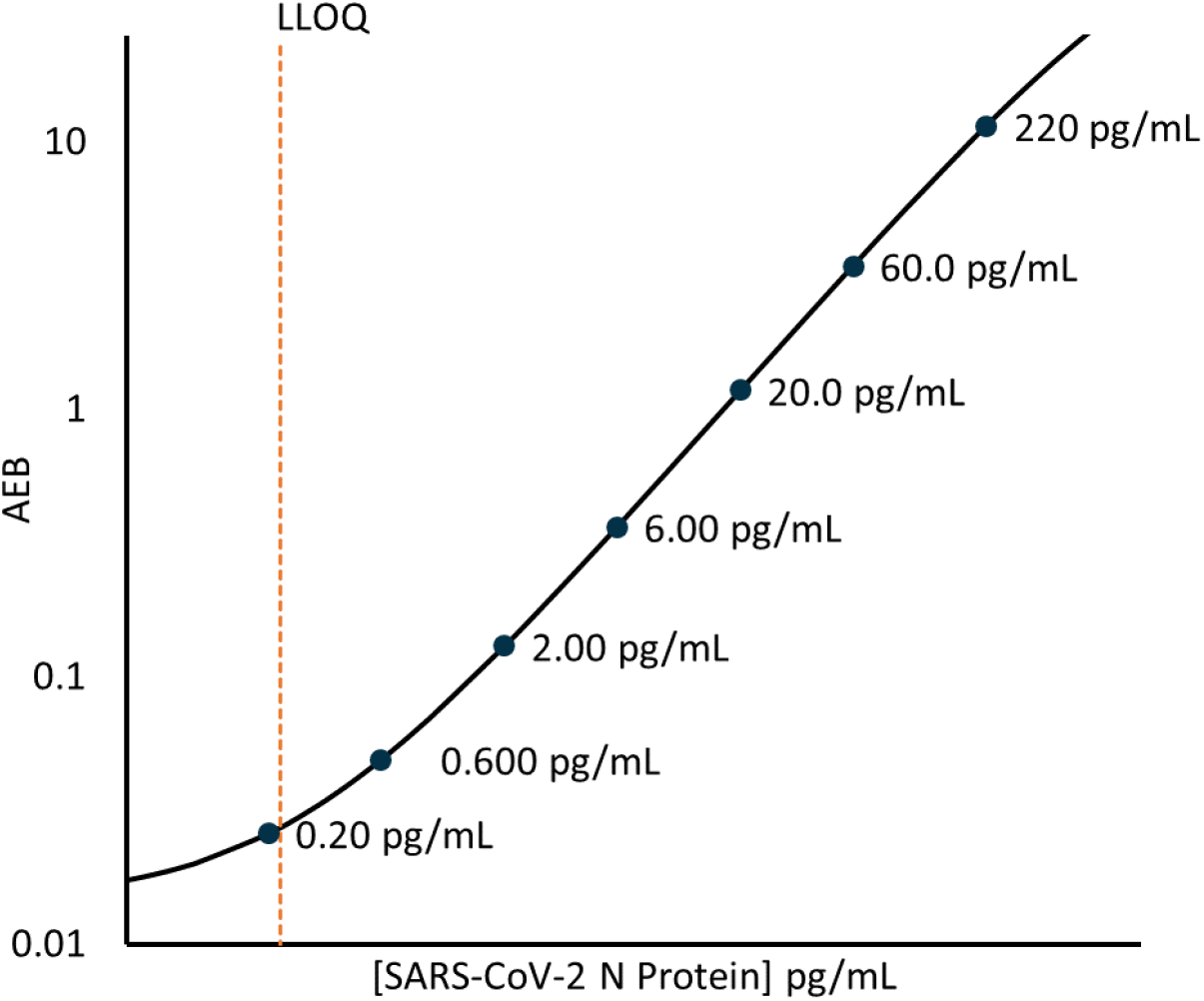
Calibration curve of the Simoa SARS-CoV-2 N Protein Advantage Assay. Lower limit of quantitation (LLoQ) is shown as the dashed line, and calibrator concentrations are denoted on graph.

## Supplemental Information

**Table S1.** Performance characteristics of Simoa SARS-CoV-2 N Protein Advantage Assay.

|  | Antigen Assay |
| --- | --- |
| Minimum Required Dilution (MRD) | 4x (serum and plasma)<br>12.5x (DBS) |
| Required Sample Volume | 25 µl (serum and plasma)<br>20 µl (DBS) |
| Assay Range (adjusted for dilution) | 0.9 – 800 pg/ml (serum and plasma)<br>2.8 – 2500 pg/ml (DBS) |
| Clinical Specificity | 100% |
| Clinical Sensitivity | 97.6% |
| Limit of Blank | 0.1 pg/ml |
| Limit of Detection | 0.32 pg/ml (0.047 TCID<sub>50</sub>/ml) |
| Limit of Quantification | 0.91 pg/ml (0.094 TCID<sub>50</sub>/ml) |
| Precision | ~6% within-run<br>~6% between-run<br>~4% between-day |
| Dilution Linearity | ~102% recovery |
| Spike Recovery | ~98% |

**Figure S2.**
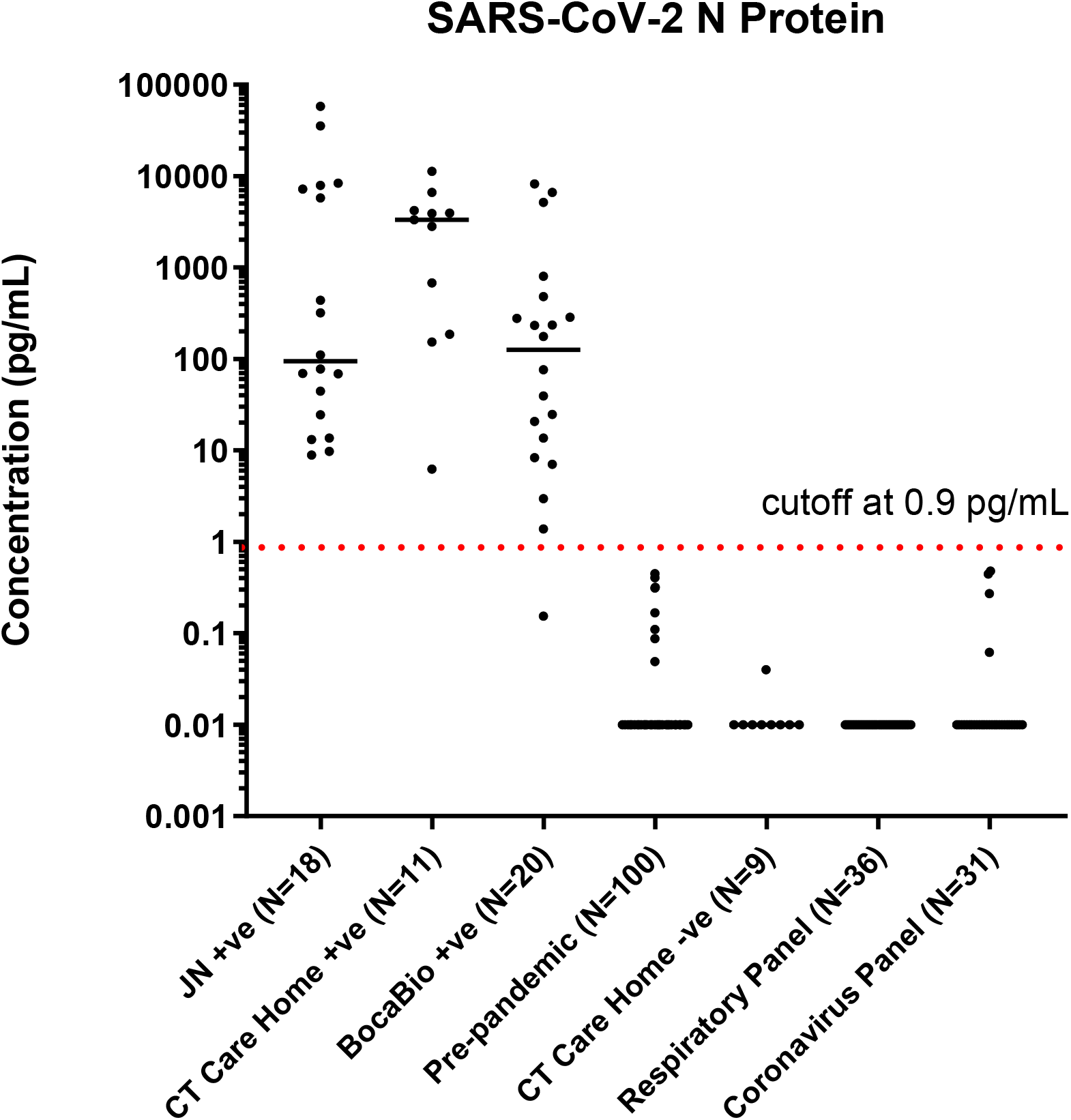
Seven sample cohorts were used to establish a preliminary clinical cutoff of 0.9 pg/ml for the SARS-CoV-2 N-Protein assay; sample numbers are shown as N in the axis label. JN +ve was K2 EDTA plasma from PCR+ donors from the U. Bonn cohort. BocaBio (+ve) were PCR+ serum from that commercial supplier. CT Care Home +ve (PCR+) and -ve (PCR-) were dried blood spot (DBS) samples from residents and staff of the care facility. Pre-pandemic samples were a mixture of serum and K2 EDTA plasma from donors acquired before December 2019 purchased from BioIVT. Respiratory Panel was K2 EDTA plasma purchased from BioIVT and from donors serologically confirmed to have been infected with combinations of H. influenza, RSV, influenza A, influenza B, parainfluenza (1-4), adenovirus, enterovirus, M. pneumoniae, Legionella, B. pertussis, and C. pneumoniae. Coronavirus Panel was serum purchased from BioIVT from donors serologically confirmed to have been infected with human coronaviruses-HKU, OC43, 229E and NL63. Preliminary cutoff of 0.9 pg/ml was chosen to confer 100% specificity over all SARS-CoV-2 negative sample cohorts.

**Figure S3.**
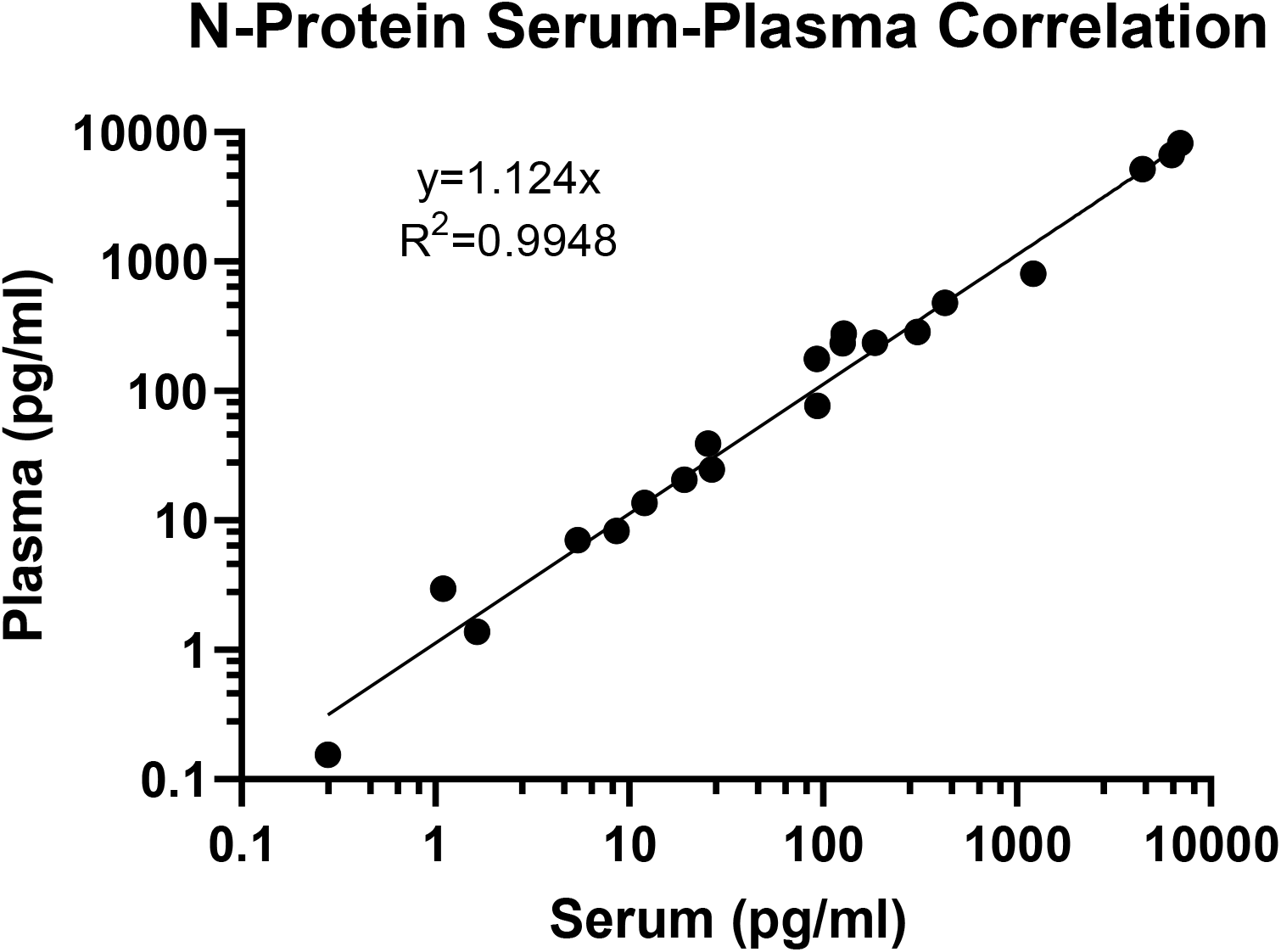
Matched serum and plasma samples from the same donors were found to have excellent correlation in N antigen levels between matrices. Twenty matched samples from BocaBio confirmed to be PCR+ were tested in both serum and K2 EDTA plasma.

**Figure S4.**
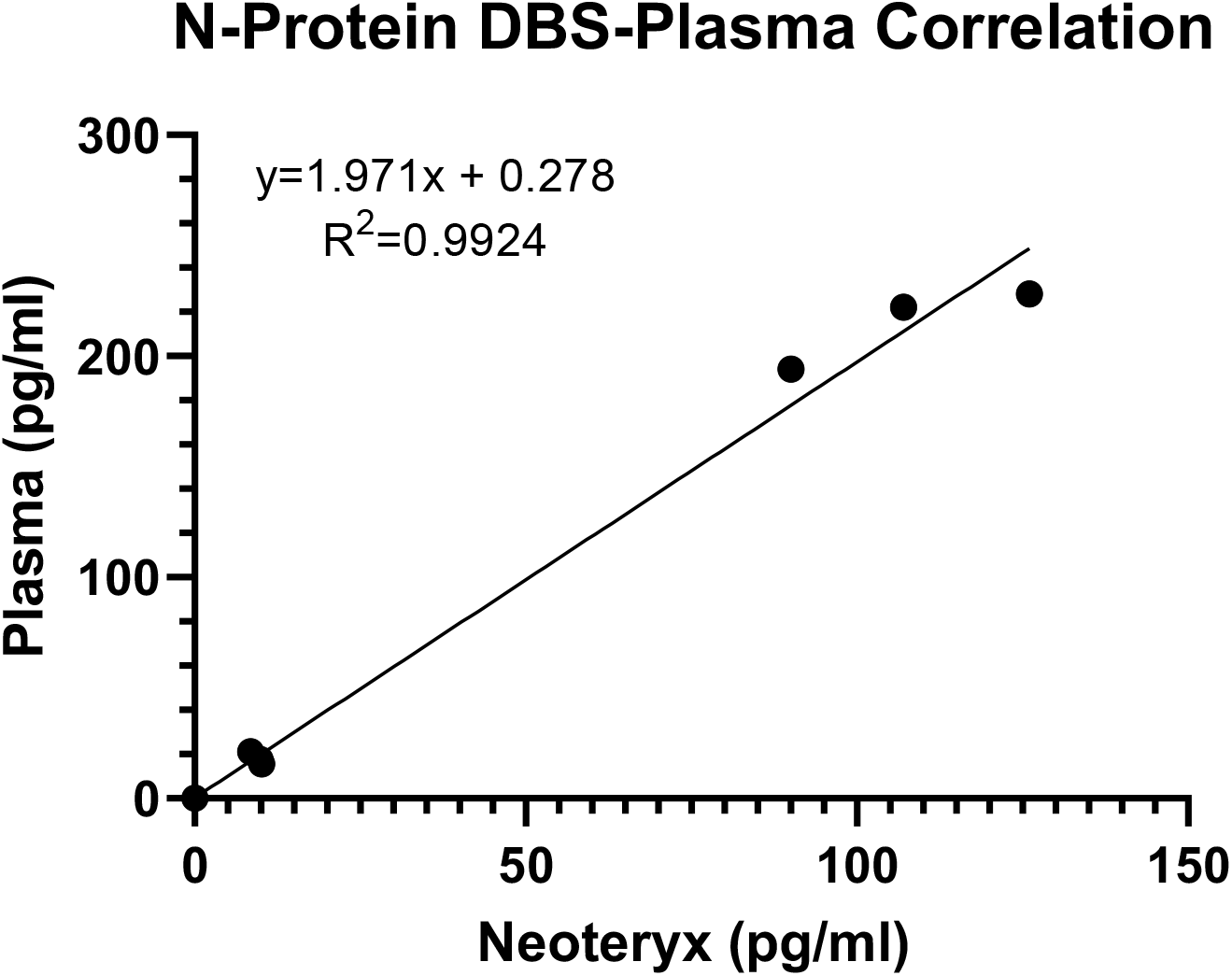
Whole blood drawn into a K2 EDTA plasma tube (3 donors) was spiked with known levels of recombinant N-protein. It was then processed into neat plasma and in parallel into Dried Blood Spots using Neoteryx Mitra tips. After extraction, both sample types were measured, showing a correlation of 0.9926. The concentration in DBS was approximately ½ of that in plasma, as expected due to the excluded volume of hematocrit which is separated from plasma.

**Figure S5.**
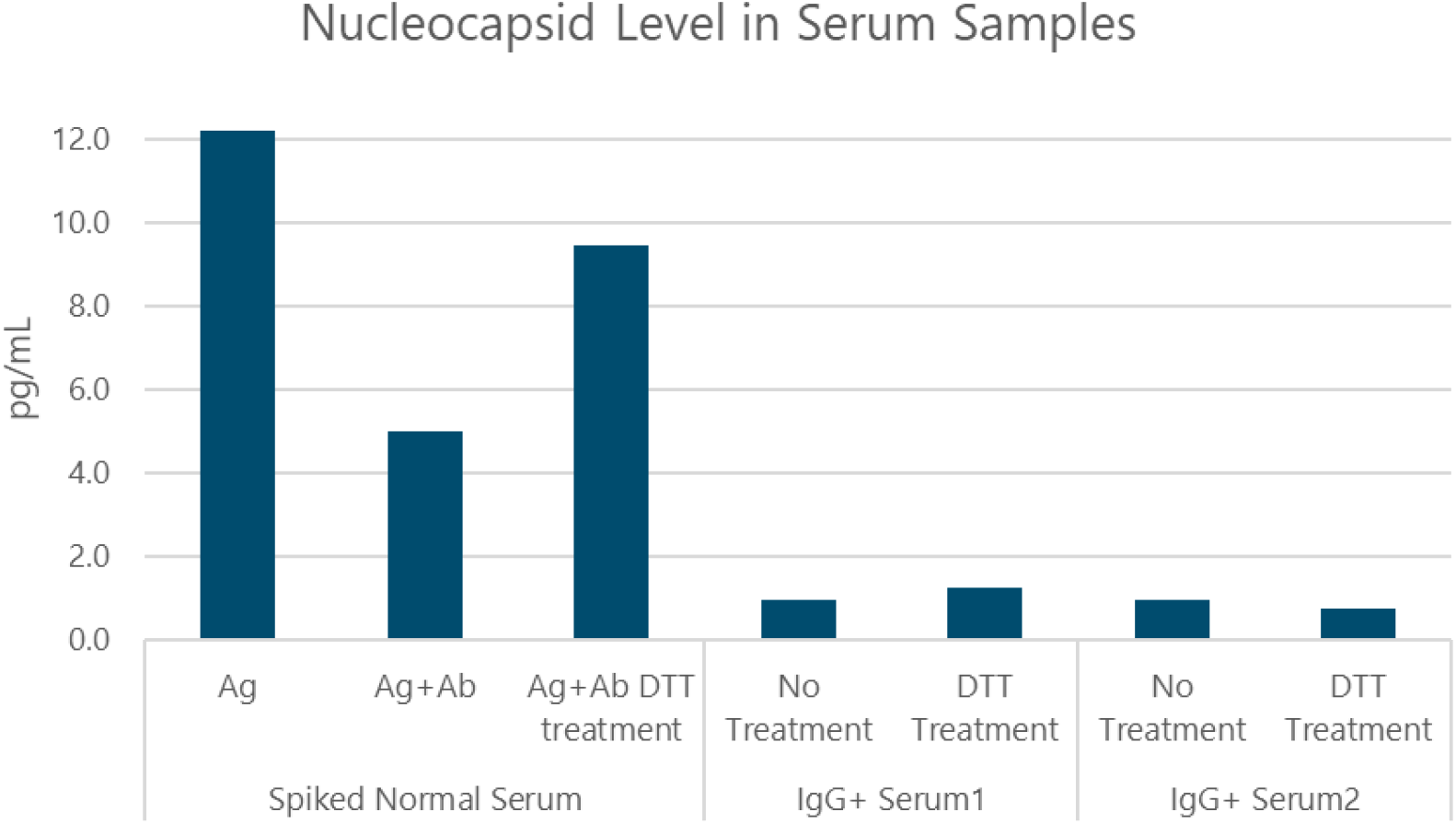
A DTT reduction protocol was established to unmask N-protein bound by antibody in serum by doing a control experiment with recombinant antigen and capture antibody spiked into sample matrix. N-protein concentration measured in serum was reduced after co-spiking with antibody, indicative of epitope masking. Adding DTT to the sample rescued 63% of the signal loss, indicating that this treatment could unmask antigen in seroconverted samples.

**Table S2.**
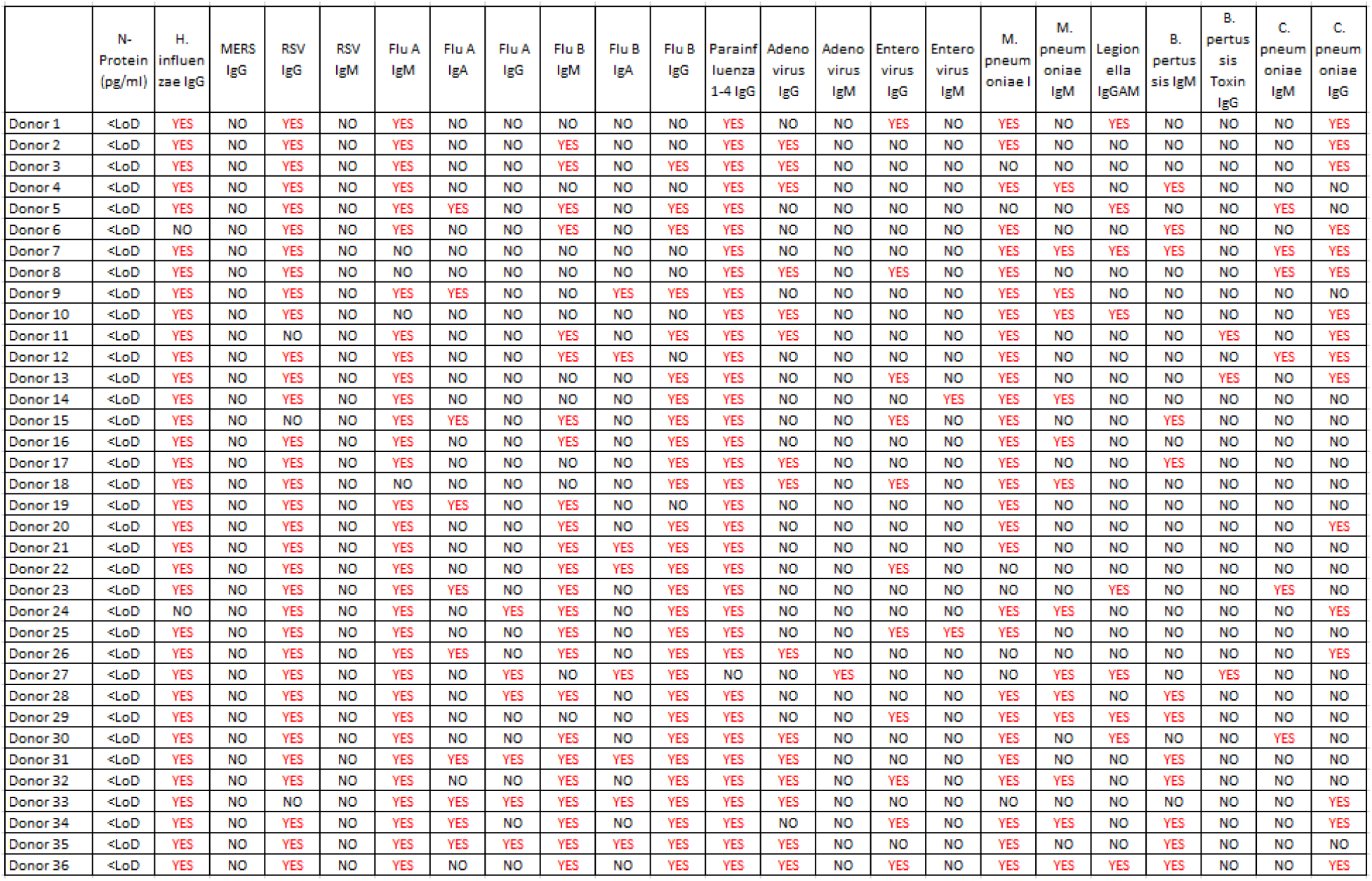
Serum from COVID-19-negative donors serologically confirmed to have been infected with common respiratory viruses, demonstrating no cross-reactivity.

|  | N-Protein<br>(pg/ml) | H.<br>influen<br>zae IgG | MERS<br>IgG | RSV<br>IgG | RSV<br>IgM | Flu A<br>IgM | Flu A<br>IgA | Flu A<br>IgG | Flu B<br>IgM | Flu B<br>IgA | Flu B<br>IgG | Parainf<br>1-4 IgG | Adeno<br>virus<br>IgG | Adeno<br>virus<br>IgM | Entero<br>virus<br>IgG | Entero<br>virus<br>IgM | M.<br>pneum<br>oniae I | M.<br>pneum<br>oniae<br>IgM | Legion<br>ella<br>IgGAM | B.<br>pertus<br>sis IgM | B.<br>pertus<br>sis Toxin<br>IgG | C.<br>pneum<br>oniae<br>IgM | C.<br>pneum<br>oniae<br>IgG |
| --- | --- | --- | --- | --- | --- | --- | --- | --- | --- | --- | --- | --- | --- | --- | --- | --- | --- | --- | --- | --- | --- | --- | --- |
| Donor 1 | <LoD | YES | NO | YES | NO | YES | NO | NO | NO | NO | NO | YES | NO | NO | YES | NO | YES | NO | YES | NO | NO | NO | YES |
| Donor 2 | <LoD | YES | NO | YES | NO | YES | NO | NO | YES | NO | NO | YES | YES | NO | NO | NO | YES | NO | NO | NO | NO | NO | YES |
| Donor 3 | <LoD | YES | NO | YES | NO | YES | NO | NO | YES | NO | YES | YES | YES | NO | NO | NO | NO | NO | NO | NO | NO | NO | YES |
| Donor 4 | <LoD | YES | NO | YES | NO | YES | NO | NO | NO | NO | NO | YES | YES | NO | NO | NO | YES | YES | NO | YES | NO | NO | NO |
| Donor 5 | <LoD | YES | NO | YES | NO | YES | YES | NO | YES | NO | YES | YES | NO | NO | NO | NO | NO | NO | YES | NO | NO | YES | NO |
| Donor 6 | <LoD | NO | NO | YES | NO | YES | NO | NO | YES | NO | YES | YES | NO | NO | NO | NO | YES | NO | NO | YES | NO | NO | YES |
| Donor 7 | <LoD | YES | NO | YES | NO | NO | NO | NO | NO | NO | NO | YES | NO | NO | NO | NO | YES | YES | YES | YES | NO | YES | YES |
| Donor 8 | <LoD | YES | NO | YES | NO | NO | NO | NO | NO | NO | NO | YES | YES | NO | YES | NO | YES | NO | NO | NO | NO | YES | YES |
| Donor 9 | <LoD | YES | NO | YES | NO | YES | YES | NO | NO | YES | YES | YES | NO | NO | NO | NO | YES | YES | NO | NO | NO | NO | NO |
| Donor 10 | <LoD | YES | NO | YES | NO | NO | NO | NO | NO | NO | NO | YES | YES | NO | NO | NO | YES | YES | YES | NO | NO | NO | YES |
| Donor 11 | <LoD | YES | NO | NO | NO | YES | NO | NO | YES | NO | YES | YES | YES | NO | NO | NO | YES | NO | NO | NO | YES | NO | YES |
| Donor 12 | <LoD | YES | NO | YES | NO | YES | NO | NO | YES | YES | NO | YES | NO | NO | NO | NO | YES | NO | NO | NO | NO | YES | YES |
| Donor 13 | <LoD | YES | NO | YES | NO | YES | NO | NO | NO | NO | YES | YES | YES | NO | YES | NO | YES | NO | NO | NO | YES | NO | YES |
| Donor 14 | <LoD | YES | NO | YES | NO | YES | NO | NO | NO | NO | YES | YES | NO | NO | NO | YES | YES | YES | NO | NO | NO | NO | NO |
| Donor 15 | <LoD | YES | NO | NO | NO | YES | YES | NO | YES | NO | YES | YES | NO | NO | YES | NO | YES | NO | NO | YES | NO | NO | NO |
| Donor 16 | <LoD | YES | NO | YES | NO | YES | NO | NO | YES | NO | YES | YES | YES | NO | NO | NO | YES | YES | NO | NO | NO | NO | NO |
| Donor 17 | <LoD | YES | NO | YES | NO | YES | NO | NO | NO | NO | YES | YES | YES | NO | NO | NO | YES | NO | NO | YES | NO | NO | NO |
| Donor 18 | <LoD | YES | NO | YES | NO | NO | NO | NO | NO | NO | YES | YES | YES | NO | YES | NO | YES | YES | NO | NO | NO | NO | NO |
| Donor 19 | <LoD | YES | NO | YES | NO | YES | YES | NO | YES | NO | NO | YES | NO | NO | NO | NO | YES | NO | NO | NO | NO | NO | NO |
| Donor 20 | <LoD | YES | NO | YES | NO | YES | NO | NO | YES | NO | YES | YES | NO | NO | NO | NO | YES | NO | NO | NO | NO | NO | YES |
| Donor 21 | <LoD | YES | NO | YES | NO | YES | NO | NO | YES | YES | YES | YES | NO | NO | NO | NO | YES | NO | NO | NO | NO | NO | NO |
| Donor 22 | <LoD | YES | NO | YES | NO | YES | NO | NO | YES | YES | YES | YES | NO | NO | YES | NO | NO | NO | NO | NO | NO | NO | NO |
| Donor 23 | <LoD | YES | NO | YES | NO | YES | YES | NO | YES | NO | YES | YES | YES | NO | NO | NO | NO | NO | YES | NO | NO | YES | NO |
| Donor 24 | <LoD | NO | NO | YES | NO | YES | NO | YES | YES | NO | YES | YES | NO | NO | NO | NO | YES | YES | NO | NO | NO | NO | YES |
| Donor 25 | <LoD | YES | NO | YES | NO | YES | NO | NO | YES | NO | YES | YES | NO | NO | YES | YES | YES | NO | NO | NO | NO | NO | NO |
| Donor 26 | <LoD | YES | NO | YES | NO | YES | YES | NO | YES | NO | YES | YES | YES | NO | NO | NO | NO | NO | NO | NO | NO | NO | YES |
| Donor 27 | <LoD | YES | NO | YES | NO | YES | NO | YES | NO | YES | YES | NO | NO | YES | NO | NO | NO | YES | YES | NO | YES | NO | NO |
| Donor 28 | <LoD | YES | NO | YES | NO | YES | NO | YES | YES | NO | YES | YES | YES | NO | NO | NO | YES | YES | NO | YES | NO | NO | NO |
| Donor 29 | <LoD | YES | NO | YES | NO | YES | NO | NO | NO | NO | YES | YES | NO | NO | YES | NO | YES | YES | YES | YES | NO | NO | NO |
| Donor 30 | <LoD | YES | NO | YES | NO | YES | NO | NO | YES | NO | YES | YES | YES | NO | NO | NO | YES | NO | YES | NO | NO | YES | NO |
| Donor 31 | <LoD | YES | NO | YES | NO | YES | YES | YES | YES | YES | YES | YES | YES | NO | NO | NO | YES | NO | NO | YES | NO | NO | NO |
| Donor 32 | <LoD | YES | NO | YES | NO | YES | NO | NO | YES | NO | YES | YES | YES | NO | YES | NO | YES | YES | NO | YES | NO | NO | NO |
| Donor 33 | <LoD | YES | NO | NO | NO | YES | YES | YES | YES | YES | YES | YES | YES | NO | NO | NO | NO | NO | NO | NO | NO | NO | YES |
| Donor 34 | <LoD | YES | NO | YES | NO | YES | YES | NO | YES | NO | YES | YES | NO | NO | YES | NO | YES | YES | NO | YES | NO | NO | YES |
| Donor 35 | <LoD | YES | NO | YES | NO | YES | YES | YES | YES | YES | YES | YES | YES | NO | NO | NO | YES | NO | NO | YES | NO | NO | NO |
| Donor 36 | <LoD | YES | NO | YES | NO | YES | NO | NO | YES | NO | YES | YES | YES | NO | YES | NO | YES | YES | YES | YES | NO | NO | YES |

**Table S3.** Serum from COVID-19-negative donors serologically confirmed to have been infected with common coronaviruses, demonstrating no cross-reactivity.

| Sample ID | SARS-CoV-2 N Protein Conc (pg/mL) | VAXARRAY CORONAVIRUS HKU + | VAXARRAY CORONAVIRUS HKU S/B RATIO (>=3.0 positive) | VAXARRAY CORONAVIRUS OC43 + | VAXARRAY CORONAVIRUS OC43 S/B RATIO (>=3.0 positive) | VAXARRAY CORONAVIRUS 229E + | VAXARRAY CORONAVIRUS 229E S/B RATIO (>=3.0 positive) | VAXARRAY CORONAVIRUS NL63 + | VAXARRAY CORONAVIRUS NL63 S/B RATIO (>=3.0 positive) |
| --- | --- | --- | --- | --- | --- | --- | --- | --- | --- |
| Donor 1 | <LoD | NO | 2.5 | NO | 1.9 | YES | 5.3 | NO | 2.7 |
| Donor 2 | <LoD | NO | 2.7 | YES | 7 | YES | 13.3 | NO | 1.4 |
| Donor 3 | <LoD | NO | 1.7 | YES | 4.2 | YES | 3.5 | NO | 2.5 |
| Donor 4 | <LoD | NO | 2.5 | NO | 2.9 | YES | 5.8 | NO | 2.1 |
| Donor 5 | <LoD | NO | 2.7 | YES | 9.8 | YES | 12.8 | NO | 2.8 |
| Donor 6 | 0.446 | YES | 4.2 | YES | 4.5 | YES | 4.5 | YES | 3 |
| Donor 7 | <LoD | YES | 4.6 | YES | 4.7 | YES | 4.7 | YES | 3.7 |
| Donor 8 | 0.272 | YES | 6.4 | YES | 5.8 | YES | 6.5 | YES | 6.5 |
| Donor 9 | <LoD | YES | 9.4 | YES | 9.5 | YES | 9.5 | YES | 9.5 |
| Donor 10 | <LoD | YES | 3.7 | YES | 3.8 | YES | 3.8 | NO | 2.1 |
| Donor 11 | <LoD | YES | 3.2 | YES | 8.8 | YES | 8.8 | YES | 3.7 |
| Donor 12 | <LoD | YES | 7.5 | YES | 10.1 | YES | 10.1 | YES | 3.1 |
| Donor 13 | <LoD | YES | 10.8 | YES | 10.9 | YES | 10.9 | YES | 10.9 |
| Donor 14 | <LoD | YES | 4.4 | YES | 4.6 | YES | 4.6 | YES | 4.6 |
| Donor 15 | <LoD | YES | 5.4 | YES | 3.3 | YES | 5.5 | NO | 2.8 |
| Donor 16 | <LoD | YES | 8.6 | YES | 11.5 | YES | 11.5 | YES | 8.4 |
| Donor 17 | 0.481 | YES | 3.1 | YES | 4.5 | YES | 5.1 | YES | 3.5 |
| Donor 18 | <LoD | YES | 6.2 | YES | 10.9 | YES | 10.9 | YES | 4.2 |
| Donor 19 | <LoD | YES | 7.9 | YES | 8 | YES | 8 | YES | 8 |
| Donor 20 | <LoD | YES | 9.3 | YES | 9.4 | YES | 9.4 | YES | 3.1 |
| Donor 21 | 0.062 | YES | 3.8 | YES | 5.1 | YES | 6.2 | YES | 6.2 |
| Donor 22 | <LoD | YES | 3.7 | YES | 5.1 | YES | 5.1 | YES | 4.8 |
| Donor 23 | <LoD | YES | 10.7 | YES | 10.8 | YES | 9.7 | YES | 4 |
| Donor 24 | <LoD | YES | 5.2 | YES | 6.5 | YES | 6.5 | YES | 5.9 |
| Donor 25 | <LoD | YES | 8.3 | YES | 8.4 | YES | 8.4 | YES | 5 |
| Donor 26 | <LoD | YES | 12.1 | YES | 15.3 | YES | 15.3 | YES | 7.5 |
| Donor 27 | <LoD | YES | 5.4 | YES | 7.8 | YES | 7.8 | YES | 5.2 |
| Donor 28 | <LoD | YES | 4.7 | YES | 4.8 | YES | 4.9 | YES | 4 |
| Donor 29 | <LoD | YES | 4.1 | YES | 4.2 | YES | 4.2 | YES | 4.2 |
| Donor 30 | <LoD | YES | 4.5 | YES | 5.4 | YES | 5.4 | YES | 5.4 |
| Donor 31 | <LoD | YES | 8.1 | YES | 12.7 | YES | 12.8 | YES | 8.4 |

**Table S4.**
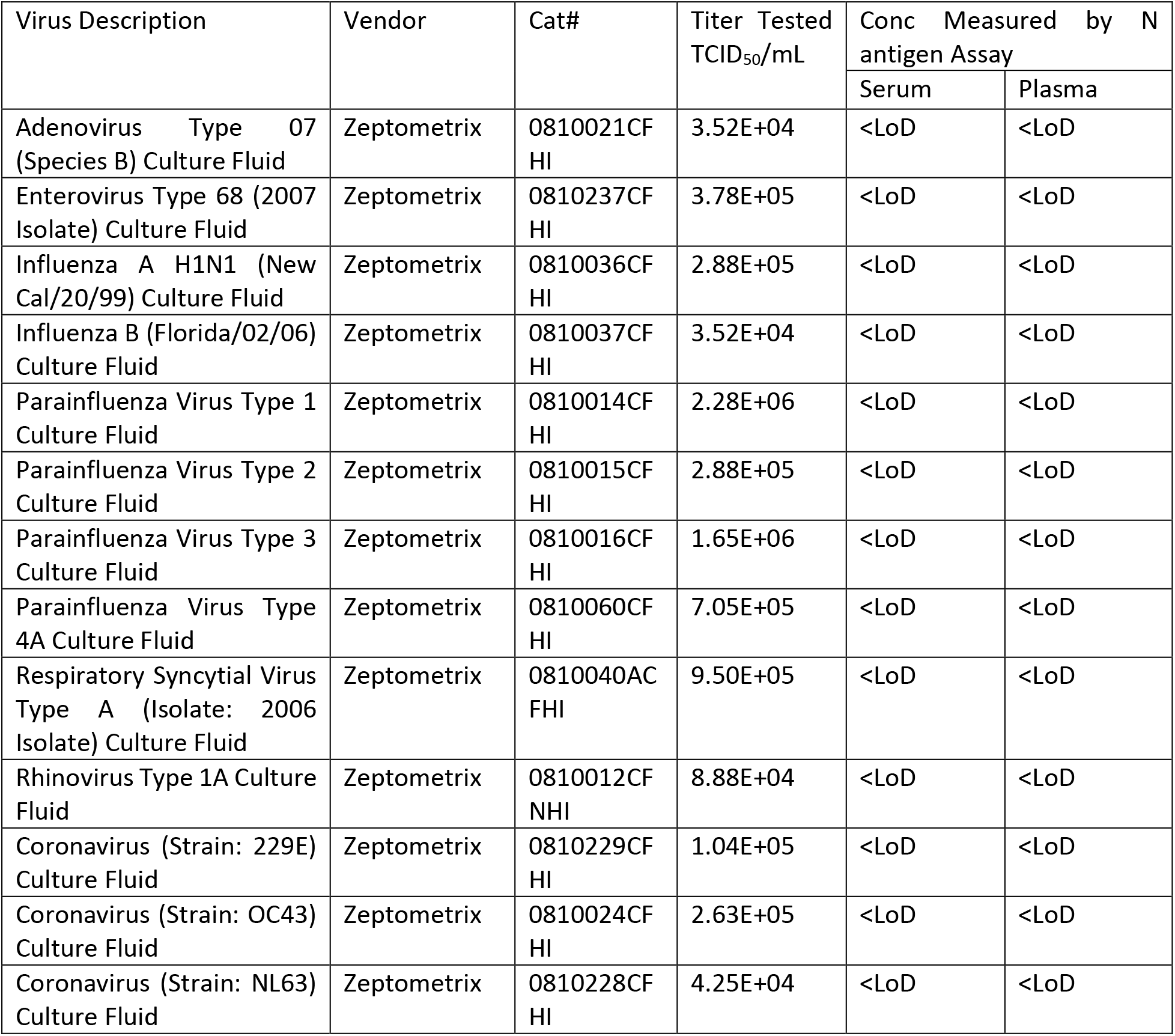
Inactivated, cultured virus was purchased from Zeptometrix, and tested for cross-reactivity at the TCID_50_ levels listed. No cross-reactivity was observed.

| Virus Description | Vendor | Cat# | Titer Tested<br>TCID<sub>50</sub>/mL | Conc Measured by N<br>antigen Assay |  |
| --- | --- | --- | --- | --- | --- |
|  |  |  |  | Serum | Plasma |
| Adenovirus Type 07<br>(Species B) Culture Fluid | Zeptomatrix | 0810021CF<br>HI | 3.52E+04 | <LoD | <LoD |
| Enterovirus Type 68 (2007<br>Isolate) Culture Fluid | Zeptomatrix | 0810237CF<br>HI | 3.78E+05 | <LoD | <LoD |
| Influenza A H1N1 (New<br>Cal/20/99) Culture Fluid | Zeptomatrix | 0810036CF<br>HI | 2.88E+05 | <LoD | <LoD |
| Influenza B (Florida/02/06)<br>Culture Fluid | Zeptomatrix | 0810037CF<br>HI | 3.52E+04 | <LoD | <LoD |
| Parainfluenza Virus Type 1<br>Culture Fluid | Zeptomatrix | 0810014CF<br>HI | 2.28E+06 | <LoD | <LoD |
| Parainfluenza Virus Type 2<br>Culture Fluid | Zeptomatrix | 0810015CF<br>HI | 2.88E+05 | <LoD | <LoD |
| Parainfluenza Virus Type 3<br>Culture Fluid | Zeptomatrix | 0810016CF<br>HI | 1.65E+06 | <LoD | <LoD |
| Parainfluenza Virus Type<br>4A Culture Fluid | Zeptomatrix | 0810060CF<br>HI | 7.05E+05 | <LoD | <LoD |
| Respiratory Syncytial Virus<br>Type A (Isolate: 2006<br>Isolate) Culture Fluid | Zeptomatrix | 0810040AC<br>FHI | 9.50E+05 | <LoD | <LoD |
| Rhinovirus Type 1A Culture<br>Fluid | Zeptomatrix | 0810012CF<br>NHI | 8.88E+04 | <LoD | <LoD |
| Coronavirus (Strain: 229E)<br>Culture Fluid | Zeptomatrix | 0810229CF<br>HI | 1.04E+05 | <LoD | <LoD |
| Coronavirus (Strain: OC43)<br>Culture Fluid | Zeptomatrix | 0810024CF<br>HI | 2.63E+05 | <LoD | <LoD |
| Coronavirus (Strain: NL63)<br>Culture Fluid | Zeptomatrix | 0810228CF<br>HI | 4.25E+04 | <LoD | <LoD |

**Table S5.** N-protein and SARS-CoV-2 specific IgG concentrations measured in the CTCH cohort for all donors and both collections.

| Donor ID | Collection One |  |  |  | Collection Two |  |  |  |
| --- | --- | --- | --- | --- | --- | --- | --- | --- |
|  | days from symptom | days from PCR | NP pg/ml | Spk IgG | days from symptom | days from last PCR | NP pg/ml | Spk IgG |
| 1 | 7 | 7 | 4226.08 | 0.06 | 14 | 14 | 4.62 | 13.41 |
| 2 | 8 | 7 | 2811.23 | 0.00 | intubated |  |  |  |
| 3 | 6 | 6 | 186.14 | 0.03 | 13 | 13 | 7.41 | 0.14 |
| 4 | 9 | 6 | 3933.59 | 0.01 | died |  |  |  |
| 5 |  | 6 | 677.47 | 0.03 |  | 13 | 925 | 0.11 |
| 6 |  | 5 | 11235.56 | 0.01 | died |  |  |  |
| 7 |  | 5 | 154.47 | 0.03 |  | 12 | 72.2 | 0.06 |
| 8 | 4 | 5 | 3349.23 | 0.01 | Opted out of study --> died |  |  |  |
| 9 |  | 5 | 3896.05 | 0.03 |  | 12 | 1051 | 0.14 |
| 11 |  | 5 | 6658.056 | 0.208 | died |  |  |  |
| 12 | -3 | 5 | 6.260 | 0.037 | 4 | 4 | 308 | 0.002 |
| 13 |  | 5 | 0.010 | 0.001 |  | 12 | 0.01 | 0.005 |
| 14 |  | 5 | 0.010 | 0.014 |  | 4 | 0.01 | 0.002 |
| 15 |  | 5 | 0.040 | 0.024 |  | 4 | 0.01 | 0.021 |
| 16 |  | 5 | 0.010 | 0.011 |  | 4 | 0.260 | 0.015 |
| 17 |  | 5 | 0.040 | 0.005 |  | 4 | 0.01 | 0.013 |
| 18 |  | 5 | 0.010 | 0.000 |  | 4 | 0.074 | 0.000 |
| 19 |  | 5 | 0.010 | 0.019 |  | 4 | 0.01 | 0.012 |
| 20 |  | 5 | 0.010 | 0.024 |  | 4 | 0.01 | 0.009 |
| 21 |  | 5 | 0.010 | 0.012 |  | 4 | 0.074 | 0.011 |
| 22 |  |  |  |  |  | 4 | 0.01 | 0.005 |
| 23 |  |  |  |  |  | 17 | 0.993 | 0.702 |
| 24 |  |  |  |  |  | 4 | 44.4 | 0.002 |
| 25 |  |  |  |  |  | 4 | 0.01 | 0.031 |
| 26 |  |  |  |  | 0 | 0 | 167 | 0.003 |
| 27 |  |  |  |  |  | 11 | 0.01 | 0.054 |
| 28 |  |  |  |  |  | 4 | 0.01 | 0.144 |

